# Applying the Nova food classification to food product databases using discriminative ingredients: a methodological proposal

**DOI:** 10.1101/2024.04.12.24305721

**Authors:** Mariana Fagundes Grilo, Beatriz Silva Nunes, Ana Clara Duran, Camila Zancheta Ricardo, Larissa Galastri Baraldi, Euridice Martinez Steele, Camila Aparecida Borges

**Affiliations:** Center for Food Studies and Research, University of Campinas, Campinas, Brazil; Department of Exercise and Nutrition Sciences, The George Washington University, Washington D.C., US; Graduate Program in Collective Health, Faculty of Medical Sciences, University of Campinas, Campinas, Brazil; Center for Epidemiological Studies in Nutrition and Health, University of São Paulo, São Paulo, Brazil; Doctoral Program in Public Health, School of Public Health, University of Chile, Santiago, Chile

**Keywords:** Ultra-processed foods, methodology, food ingredients, classification, food additives, food substances

## Abstract

**Background:** Growing interest in the Nova food classification system surged among various stakeholders, driven primarily by compelling evidence linking the consumption of ultra-processed foods (UPF) to negative health outcomes. However, a more standardized approach could incentivize the identification of UPF in regulatory processes.

**Objective:** To propose replicable methods to identify UPF that, by testing the sensitivity and specificity of these methods using a large sample of packaged foods from the 2017 Brazilian Food Labels Database.

**Methods:** We created five scenarios to identify UPF using food substances and food additives typically found in UPF and compared them with the most frequently employed Nova food classification process based on the product name and food categories, considered the ‘classic method’ to identify UPF. We estimated the proportion of foods and beverages identified as UPF using different scenarios based on the presence of discriminative ingredients. We used a diagnostic test and a receiver operating characteristic (ROC) to understand which of the five scenarios performed better compared to the ‘classic method’ to identify UPF. Finally, we conducted a sensitivity analysis to test the role of vitamins and minerals in identifying UPF.

**Results:** We found variations in UPF prevalence from 47% to 72% across scenarios, compared to 70% using the ‘classic method’ to identify UPF in Brazilian packaged foods. Despite its cautious approach, the scenario using additives of a sole cosmetic function and food byproducts (scenario 3) identified a 65% UPF, while maintaining reasonable sensitivity and specificity, and the best-performing ROC curve. There was no significant difference in identifying UPF when comparing the addition of vitamins and minerals to the food additives with sole cosmetic function.

**Conclusion:** Our study sheds light on the importance of detailed criteria to identify UPF and offers the research community a standardized method to identify UPF.

## Introduction

The Nova food classification system has, over the past two decades, shifted the focus of dietary intake and food composition assessment research from nutrient composition to including the extent and purpose of industrial processing of foods and beverages (1). This system classifies foods into 4 major groups: unprocessed and minimally processed foods, processed culinary ingredients, processed foods, and ultra-processed foods (UPF). The latter are defined as industrial formulations, typically made of food components that have been either modified or recombined, with little or no whole foods, as well as industrial substances and food additives aimed at increasing durability and enhancing or modifying sensory characteristics such as color, taste, odor and texture of foods (2). The UPF are developed to be palatable, cheaper, and designed to have a strong market appeal. Additionally, they are commonly ready to eat, heat, or be reconstituted, often requiring little to no culinary preparation, such as packaged fruit juice concentrates, breakfast cereals, cookies, and many ready-to-eat packaged meals, such as frozen lasagna and nuggets (2).

Over the years, the Nova food classification has proved to be efficient in predicting the nutritional quality of diets and in identifying the increasing consumption of UPF over time in diverse populations around the world (3-7). The Nova food classification has been applied to observational studies (8, 9), cohort studies (10-12), and randomized trials (13) to assess the link between UPF consumption and health outcomes. At the moment, evidence is available on the associations of UPF with health outcomes such as weight gain (14), type 2 diabetes (15), cardiometabolic diseases (16), cerebrovascular disease (16), cancer (17), premature deaths (18), all-cause mortality (16), among others (9, 19, 20).

To discourage or curb rising UPF consumption, governments across the globe enacted policies to regulate their distribution, promotion, and purchase, such as mandatory front-of-pack nutrition labeling, marketing and procurement restrictions, and taxation (21). Over the last years, the criteria for regulating foods and beverages have been refined. However, for UPF to be targeted, policies require the clear identification of these foods. While some strategies to identify UPF based on food name, food category or food description (22, 23) have been suggested when detailed food composition information is lacking, a more standardized approach based on food composition information is needed for policies that require zero to very low uncertainty in the identification of targeted food and beverage items.

The latest version of the Nova food classification method published in 2019 mentions that a practical way to identify UPF is by checking if the list of ingredients (when available) contains at least one item that characterizes UPF, which includes “food substances never or rarely used in kitchens (such as high-fructose corn syrup, hydrogenated or interesterified oils, and hydrolyzed proteins), or classes of additives designed to make the final product palatable or more appealing (such as flavors, flavor enhancers, food colorings, emulsifiers, emulsifying salts, sweeteners, thickeners, and anti-foaming, bulking, carbonating, foaming, gelling, and glazing agents)” (2). More recently, UPF identifying methods using food additives of ‘cosmetic use’ along with the presence of high levels of nutrients of public health concern (e.g. sugar, sodium, and saturated fat) have been proposed (24, 25), advancing the discussion on optimum UPF identification criteria for policy design.

This recent literature proposes norms to deal with the food additives with varying nomenclatures and functions as well as the role of the presence of high levels of nutrients of public health concern that are usually found in UPF. Our study enhances the evolving literature by proposing replicable methods to identify UPF that, by testing the sensitivity and specificity of these methods using a large sample of packaged foods from the 2017 Brazilian Food Labels Database, aims to reduce classification uncertainty.

## Methods

### i. Data source

In this cross-sectional study, we used data from the 2017 Brazilian Food Labels Database that includes a large sample of foods and beverages sold by the five major food retailers in Brazil (26). To the best of our knowledge, this is the most comprehensive database with detailed information on the food composition of packaged foods sold in Brazil (26). Detailed information on how the data were collected is available elsewhere (26). Shortly, data collection was held in 2017 and took place in stores located in low- and high-income neighborhoods of Sao Paulo, Brazil’s largest city. Because one of the top five food retailers was only present in the northeast of the country, data collection was also held in Salvador, their largest market.

The 2017 Brazilian Food Labels Database contains 11,434 foods and beverages. Because we wanted to use the list of ingredients to classify foods and beverages according to the Nova classification, foods and beverages that did not provide the list of ingredients (n=1,574) on their package were excluded, totaling 9,860 unique products. It is important to note the in the Brazilian legislation, products that contain only one ingredient are exempted to show the list of ingredients on the package, i.e., in this case, the products that were excluded were probably fresh (e.g. rice beans, fresh fruits and vegetables) or culinary ingredient with only one ingredient (e.g. sugar, salt, oil).

For the purpose of our study, we created five UPF identification criteria, according to the latest Nova food classification published in 2019 that says that UPF can be identified by the presence of food substances never or rarely used in kitchens and certain food additives. The food substances never or rarely used in kitchens (hereafter referred to as ‘food byproduct’, i.e., any material that is produced as a secondary result during the manufacturing or production process of food items), include high-fructose corn syrup, hydrogenated or interesterified oils, and hydrolyzed proteins (2). For food byproduct, we included added sugars, carbohydrates, modified oils, protein and fiber sources based on the definitions proposed by Zancheta et al., (2) (Supplementary file 3).

The food additives that characterize UPF are the ones that can modify sensory characteristics of the products, like “colorants, flavors, artificial sweeteners, emulsifiers” (hereafter referred to as ‘food additives with cosmetic function’) (1). To identify and classify food additives we used the information available in the 2023 Codex Alimentarius (2), and translated the names of food additives according to the National Health Surveillance Agency (ANVISA) (27). Because food additives can have more than one function, we identified and distinguished food additives with sole a cosmetic function from food additives that could also have other functions.

In agreement with the latest proposed definition of UPF (1), we considered food additives with cosmetic functions the following: flavors, flavor enhancers, food colorings, emulsifiers, emulsifying salts, sweeteners, thickeners, and anti-foaming, bulking, carbonating, foaming, gelling and glazing agents (2) (Supplementary file 1). Besides looking for food additives that serve a cosmetic function in the list of ingredients, we also checked for non-technical terms for flavorings, such as ‘natural flavoring,’ as referenced in Brazilian legislation (RDC No. 2) (Supplementary file 2). According to this regulation, it is not required to list each specific substance that makes up the flavorings on the ingredient list. Instead, the label can use general terms such as ‘contains flavoring (28).

The five proposed scenarios and the classic classification method we are comparing them with (the most frequently used method to identify UPF using food names and categories which we defined as the ‘classic method’ to identify UPF) are described below:

<u>Scenario 1:</u> Presence of at least one food byproduct.

<u>Scenario 2:</u> Presence of at least one food additive with sole cosmetic function.

<u>Scenario 3:</u> Presence of at least one food byproduct or a food additive with sole cosmetic function.

<u>Scenario 4:</u> Presence of at least one food additive that can serve as an additive with cosmetic function.

<u>Scenario 5:</u> Presence of at least one food byproduct or a food additive that can serve as an additive of cosmetic function.

<u>‘Classic method’ to identify UPF:</u> Frequently used method to identify UPF by food names and food categories.

All terms used to identify food additives and food byproducts in the five proposed scenarios are available in Supplementary file 4.

In addition, because vitamins and minerals can be added to foods and beverages with the function of preservatives, in fortification efforts, but also to enhance sensory aspects such as colorings, they pose a challenge of its own. In the latter instance, they may be indicative of ultra-processing (29). Thus, given their multifaceted functions, we conducted a sensitivity analysis comparing scenario 2 (identification of UPF through at least one food additive with sole cosmetic function) with a similar scenario that included vitamins and minerals (described in Supplementary File 5). We also compared scenario 3 (identification of UPF through at least one food byproduct and food additive with sole cosmetic function) with a similar scenario that included vitamins and minerals.

### ii. Food categories

To simplify the analytical process, foods and beverages available in the Brazilian Food and Beverage Labels Database were categorized into the 25 food categories described in previous studies (26, 30): Breakfast cereals and granola bars; Bakery products; Convenience foods; Unsweetened dairy products; Sweetened dairy products; Salty snacks; Cookies; Canned vegetables; Oils and fats; Sauces and dressings; Coffee and tea; Candies and desserts; Cereals, beans, other grain products; Packaged fruits and vegetables; Meat, poultry, seafood, and egg; Sugar and other noncaloric sweeteners; Processed meats; Juices; Nectars; Fruit-flavored drinks; Soda; Other beverages; Nuts and seeds; Cheeses; Fruit preserves.

### iii. Statistical Analysis

We estimated the prevalence of UPF in the Brazilian Food Labels Database using the five proposed scenarios and the ‘classic method’ to identify UPF, overall and by food category (29). Subsequently, we conducted diagnostic tests to assess the sensitivity, specificity, positive predictive value, and negative predictive value for each scenario in comparison with the ‘classic method’ to identify UPF. In addition, we developed the receiver operating characteristic (ROC) curve to evaluate the performance of the scenarios. The ROC curve is a graphical representation of a binary classifier’s performance, plotted by sensitivity (true positive rate) against 1 – specificity (false positive rate). Its effectiveness is primarily gauged by the Area Under the Curve (AUC), with values closer to 1 indicating better performance. A superior model’s ROC curve approaches the top-left corner, reflecting a high sensitivity without a significant increase in false positives. The curve’s initial steepness and its concave shape towards the top-left are also signs of a robust classifier, indicating an effective balance between sensitivity and specificity. In essence, the closer and more bowed towards the top-left the curve is, the better the model is at distinguishing between the two classes. Finally, in the sensitivity analysis, we assessed the proportion of UPF identified, including vitamins and minerals, in scenarios 2 and 3. We compared these proportions among the scenarios using a proportion test and repeated the diagnostic test (sensitivity, specificity, positive predictive value, and negative predictive value).

Analyses were performed with Stata/MP 16.1, College Station, TX: StataCorp LLC.

## Results

The prevalence of UPF in the Brazilian food supply across the five scenarios ranged from 47.1% in scenario 1 to 71.7% in scenario 5, compared with 70.5% using the ‘Classic method’ to identify UPF (Figure 1).

**Figure 1.**
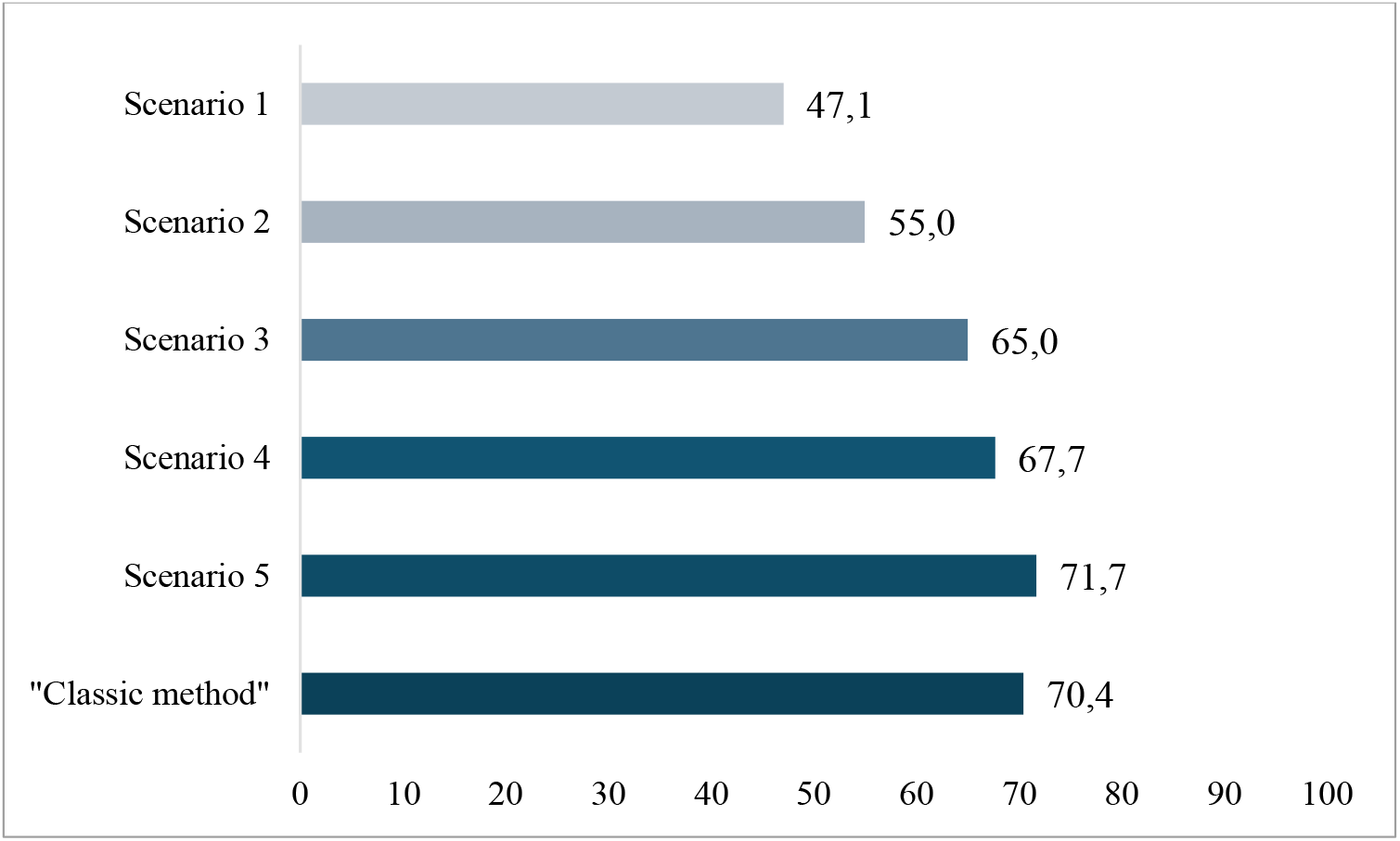
Prevalence of ultra-processed foods in the Brazilian food supply across the six proposed scenarios and the ‘classic method’ to identify ultra-processed foods (UPF) Brazilian Food Labels Database, 2017. *Scenario 1: Presence of at least one food byproduct; Scenario 2: Presence of at least one food additive with sole cosmetic function; Scenario 3: Presence of at least one food byproduct or a food additive with sole cosmetic function; Scenario 4: Presence of at least one food additive that can serve as an additive with cosmetic function; Scenario 5: Presence of at least one food byproduct or a food additive that can serve as an additive of cosmetic function; ‘Classic method’ to identify UPF: Frequently used method to identify UPF by food names and food categories.

**Figure 2.**
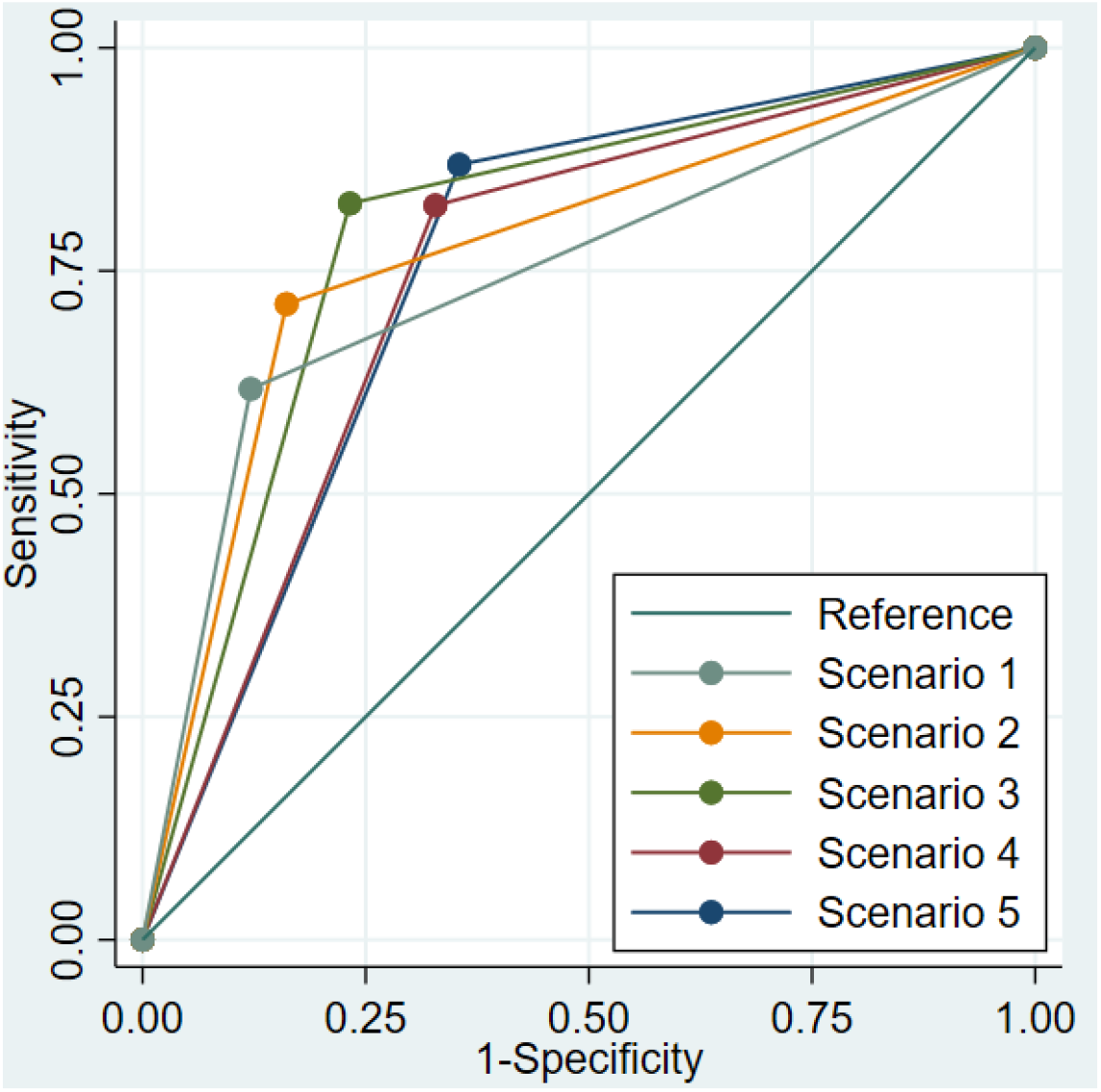
ROC curve for the six proposed scenarios having the ‘classic method’ to identify ultra-processed foods (UPF) as the reference. *Scenario 1: Presence of at least one food byproduct; Scenario 2: Presence of at least one food additive with sole cosmetic function; Scenario 3: Presence of at least one food byproduct or a food additive with sole cosmetic function; Scenario 4: Presence of at least one food additive that can serve as an additive with cosmetic function; Scenario 5: Presence of at least one food byproduct or a food additive that can serve as an additive of cosmetic function; ‘Classic method’ to identify UPF: Frequently used method to identify UPF by food names and food categories.

Table 1 shows that some food categories are consistently categorized as UPF in all scenarios, such as bakery products, sweetened dairy products, cookies, candies and desserts. However, other food categories have greater variability depending on the scenario, i.e. the UPF identification criterion, for example among unsweetened dairy products, canned vegetables, sauces and dressings, coffee and tea.

**Table 1.** Prevalence of ultra-processed foods (UPF) by scenarios of UPF identification by food category. Brazilian Food Labels Database, 2017.

| Food category | Total | Scenario 1 |  | Scenario 2 |  | Scenario 3 |  | Scenario 4 |  | Scenario 5 |  | 'Classic method' |  |
| --- | --- | --- | --- | --- | --- | --- | --- | --- | --- | --- | --- | --- | --- |
|  |  | UPF |  | UPF |  | UPF |  | UPF |  | UPF |  | UPF |  |
|  |  | n | % | n | % | n | % | n | % | n | % | n | % |
| Breakfast cereals and granola bars | 308 | 233 | 75.6 | 230 | 74.7 | 270 | 87.7 | 285 | 92.5 | 294 | 95.5 | 308 | 100.0 |
| Bakery products | 594 | 431 | 72.6 | 304 | 51.2 | 486 | 81.8 | 451 | 75.9 | 509 | 85.7 | 594 | 100.0 |
| Convenience foods | 795 | 390 | 49.1 | 447 | 56.2 | 549 | 69.1 | 514 | 64.7 | 586 | 73.7 | 795 | 100.0 |
| Unsweetened dairy products | 174 | 6 | 3.4 | 0 | 0.0 | 6 | 3.4 | 49 | 28.2 | 50 | 28.7 | 2 | 1.1 |
| Sweetened dairy products | 483 | 459 | 95.0 | 432 | 89.4 | 476 | 98.6 | 465 | 96.3 | 476 | 98.6 | 483 | 100.0 |
| Salty snacks | 356 | 140 | 39.3 | 239 | 67.1 | 257 | 72.2 | 261 | 73.3 | 272 | 76.4 | 346 | 97.2 |
| Cookies | 747 | 524 | 70.1 | 589 | 78.8 | 660 | 88.4 | 665 | 89.0 | 682 | 91.3 | 747 | 100.0 |
| Canned vegetables | 345 | 8 | 2.3 | 25 | 7.2 | 29 | 8.4 | 44 | 12.8 | 48 | 13.9 | 0 | 0.0 |
| Oils and fats | 294 | 23 | 7.8 | 68 | 23.1 | 73 | 24.8 | 100 | 34.0 | 103 | 35.0 | 46 | 15.6 |
| Sauces and dressings | 791 | 286 | 36.2 | 427 | 54.0 | 489 | 61.8 | 500 | 63.2 | 542 | 68.5 | 751 | 94.9 |
| Coffee and tea | 68 | 0 | 0.0 | 30 | 44.1 | 30 | 44.1 | 30 | 44.1 | 30 | 44.1 | 5 | 7.4 |
| Candies and desserts | 1,218 | 960 | 78.8 | 998 | 81.9 | 1144 | 93.9 | 1,108 | 91.0 | 1,167 | 95.8 | 1,210 | 99.3 |
| Cereals, beans, other grain products | 463 | 25 | 5.4 | 73 | 15.8 | 91 | 19.7 | 108 | 23.3 | 119 | 25.7 | 62 | 13.4 |
| Packaged fruits and vegetables | 299 | 0 | 0.0 | 4 | 1.3 | 4 | 1.3 | 4 | 1.3 | 4 | 1.3 | 0 | 0.0 |
| Meat, poultry, seafood, and egg | 49 | 0 | 0.0 | 0 | 0.0 | 0 | 0.0 | 3 | 6.1 | 3 | 6.1 | 0 | 0.0 |
| Sugar and other noncaloric sweeteners | 66 | 14 | 21.2 | 37 | 56.1 | 41 | 62.1 | 42 | 63.6 | 42 | 63.6 | 44 | 66.7 |
| Processed meats | 810 | 381 | 47.0 | 530 | 65.4 | 572 | 70.6 | 577 | 71.2 | 594 | 73.3 | 617 | 76.2 |
| Juices | 144 | 41 | 28.5 | 34 | 23.6 | 55 | 38.2 | 36 | 25.0 | 55 | 38.2 | 46 | 31.9 |
| Nectars | 160 | 92 | 57.5 | 131 | 81.9 | 137 | 85.6 | 140 | 87.5 | 143 | 89.4 | 160 | 100.0 |
| Fruit-flavored drinks | 220 | 138 | 62.7 | 214 | 97.3 | 216 | 98.2 | 217 | 98.6 | 218 | 99.1 | 220 | 100.0 |
| Sodas | 106 | 26 | 24.5 | 104 | 98.1 | 104 | 98.1 | 105 | 99.1 | 105 | 99.1 | 106 | 100.0 |
| Other beverages | 286 | 141 | 49.3 | 219 | 76.6 | 238 | 83.2 | 249 | 87.1 | 259 | 90.6 | 269 | 94.1 |
| Nuts and seeds | 72 | 8 | 11.1 | 8 | 11.1 | 12 | 16.7 | 8 | 11.1 | 12 | 16.7 | 8 | 11.1 |
| Cheeses | 607 | 109 | 18.0 | 174 | 28.7 | 241 | 39.7 | 500 | 82.4 | 509 | 83.9 | 126 | 20.8 |
| Fruit preserve | 405 | 208 | 51.4 | 104 | 25.7 | 231 | 57.0 | 214 | 52.8 | 248 | 61.2 | 1 | 0.2 |

\*Scenario 1: Presence of at least one food byproduct; Scenario 2: Presence of at least one food additive with sole cosmetic function; Scenario 3: Presence of at least one food byproduct or a food additive with sole cosmetic function; Scenario 4: Presence of at least one food additive that can serve as an additive with cosmetic function; Scenario 5: Presence of at least one food byproduct or a food additive that can serve as an additive of cosmetic function; ‘Classic method’ to identify UPF: Frequently used method to identify UPF by food names and food categories.

Using a diagnostic test to compare the five scenarios of UPF identification with the ‘classic method’ to identify UPF, we found increased sensitivity from scenario 1 (61.8%) to scenario 5 (86.9%), and decreased specificity (from 87.9% in scenario 1 to 64.5% in scenario 6) (Table 2).

**Table 2.** Diagnostic tests comparing the identification of ultra-processed foods (UPF) using the 6 scenarios and ‘classic method’. Brazilian Food Labels Database, 2017.

| Statistical metrics | Scenario 1 | Scenario 2 | Scenario 3 | Scenario 4 | Scenario 5 |
| --- | --- | --- | --- | --- | --- |
| Sensitivity | 61.8% | 71.3% | 82.6% | 82.3% | 86.9% |
| Specificity | 87.9% | 83.9% | 76.8% | 67.2% | 64.5% |
| Positive predictive value | 92.4% | 91.3% | 89.4% | 85.7% | 85.4% |
| Negative predictive value | 49.1% | 55.1% | 64.9% | 61.5% | 67.4% |

\*Scenario 1: Presence of at least one food byproduct; Scenario 2: Presence of at least one food additive with sole cosmetic function; Scenario 3: Presence of at least one food byproduct or a food additive with sole cosmetic function; Scenario 4: Presence of at least one food additive that can serve as an additive with cosmetic function; Scenario 5: Presence of at least one food byproduct or a food additive that can serve as an additive of cosmetic function; ‘Classic method’ to identify UPF: Frequently used method to identify UPF by food names and food categories.

We observe that the area under the curve (AUC) was 0.748 for scenario 1, 0.776 for scenario 2, 0.797 for scenario 3, 0.748 for scenario 4 and 0.757 for scenario 5. Scenario 3 had the highest AUC, indicating better performance compared to the “classic method” used as a reference for identifying UPF.

Noteworthy, in the sensitivity test, the addition of vitamins and minerals that can or cannot serve as food additives with a cosmetic function in the scenarios with food additives with sole cosmetic function to identify UPF (Supplementary file 6) showed no statistical differences as much as taking into account the presence of food byproducts or food additives that can serve as food additives with a cosmetic function (scenarios 3 and 6) or the prevalence of UPF.

## Discussion

In this study, we found that the choice of food classification method can significantly influence the estimated prevalence of UPF in the Brazilian food supply. The prevalence varied by approximately 25 percentage points among five different scenarios that included one or more UPF known markers, compared with a prevalence rate of 70% when we deployed a frequently used UPF identification criteria that considers only the name and category of each food or beverage. Additionally, we identified a scenario that successfully captured a high proportion of UPF while maintaining satisfactory results in classification according to the ROC curve when using the ‘classic method’ as the classifier. Scenario 3, which included the presence of at least one food byproduct or a food additive with sole cosmetic function, demonstrated effectiveness in capturing UPF.

The comparison between scenarios could aid future studies employing the Nova classification and assist in shaping public policies that need to distinguish UPF from other foods and beverages. In view of the urgency of having standardized and replicable methods to identify UPF, other studies have used the list of ingredients with this purpose, especially to assess UPF consumption (29), and more recently to identify the presence of nutrients of public health concern in the food supply (24). In our case, the reliance on discriminatory ingredients, such as a combination of food byproducts and food additives, introduces a level of detail and complexity in the classification process that adds to the literature on how to best operationalize the UPF construct to be used in policies that require no or very low uncertainty in the definition of UPF. The fact that Scenario 3 aligned more closely with the ‘classic method’ to identify UPF suggests that this scenario may provide more accurate UPF estimates.

Although the list of ingredients does not provide information about the food matrix, using discriminatory ingredients that correspond to foods byproducts and food additives with the function of altering the food or beverage’s physical characteristic allows for a more systematized method to identify UPF (2), and can help evaluators to reach a consensus on the identification of these foods (31).

Regarding food categories, our findings showed that certain categories are more likely to consist solely of UPF, like sweetened dairy products and candies, demonstrating that, although the chance of misclassification using the classic method (which only includes the name and category of foods) is low for some food categories. In fact, another study showed that 100% of sweet cookies, savory biscuits, margarine, cakes, sweet pies, chocolate, dairy beverages and ice cream classified as UPF in the same the Brazilian database of foods and beverages contained at least one additive with a cosmetic function or an excess of certain nutrients of interest to public health, such as sugars, salt, oils and fats (24).

For other food categories, such as convenience foods and unsweetened dairy products, the chance of misclassification was considerable high. These results reinforce the existence of higher variation in the ingredient composition and suggest that auxiliary or alternative methods should be employed for certain categories. In the case of coffee and tea, the prevalence of UPF was the same across all 6 scenarios (44.1%), but differed from the ‘classic method’ to identify UPF (7.4%), suggesting that the ‘classic method’ to identify UPF may underestimate the prevalence of UPF in this beverage category. For instance, they may be considered fresh or minimally processed by name and food group, but they may contain markers of ultra-processing in the list of ingredients, such as additives with cosmetic function.

The trade-off between sensitivity and specificity in the diagnostic test comparing all the five scenarios with the ‘classic method’ to identify UPF, while an increase in sensitivity from scenario 1 to scenario 5 is promising in accurately identifying true positive cases (decreasing the false negative UPF), there is an expected decrease in specificity, due to increasing false positive UPF. We highlight that even with a more conservative approach to identify UPF, scenario 3 demonstrated a sensitivity of 82.6% and a specificity of 76.8%. This means that using the criteria of at least one food additive with sole cosmetic function, or a food byproduct to identify UPF, resulted in high sensitivity, while maintaining an acceptable level of specificity.

Finally, vitamins and minerals can be added to foods for fortification, aiming to improve the nutritional profile and address deficiencies. However, the same food additives can also be used for cosmetic functions, such as enhancing color, texture, or shelf life, without providing significant nutritional benefits. The presence of vitamins and minerals in a food product doesn’t clearly indicate whether the primary intent is fortification or cosmetic function, creating ambiguity in classification. Additionally, food labels often do not distinguish between the functional purposes of added vitamins and minerals. Because these factors complicate efforts to accurately identify and classify UPF, we conducted a sensitivity analysis and we found that adding vitamins and minerals that can or cannot serve as food additives with a cosmetic function in the scenarios with food additives of sole cosmetic function to identify UPF did not change the identification of UPF. In other words, the presence of vitamins and minerals, regardless of their function, did not impact the classification of the foods as UPF and suggest that future research could focus more on the core characteristics of UPF.

The present study is not free of limitations that should considered in the interpretation of our findings. First, we need to consider that some food additives may have more than one function in foods and beverages (32). To address this, we proposed different scenarios, including those that identify food additives with a sole cosmetic function. Second, caution in the reproduction of our proposed scenarios in other contexts should be exerted. For instance, local food and labeling legislation regarding the listing of ingredients in packages should be considered when one decides whether other terms for food additives should be included.

Specifically, Brazilian food legislation allows companies to use the term ‘flavorings’ to denote the presence of food flavorings that may not be explicitly listed in the Codex Alimentarius. Researchers and policymakers in other countries need to adapt the proposed scenarios to fit their local food labeling laws, considering whether certain terms can obscure the presence of specific additives and thus affect UPF classification. Despite these differences, it is widely recognized that UPFs have a similar ingredient composition worldwide (24), so while local adaptations are necessary, the core principles of UPF identification should remain consistent.

Another important aspect to consider in the Brazilian context is that it is not mandatory to list the composition of compound ingredients on food labels. This lack of specificity can lead to an underestimation of the presence of cosmetic additives in foods and beverages. If more detailed information were available, it’s likely that a greater number of products would be classified as UPF, due to the higher apparent degree of processing. Additionally, this step-by-step approach to identifying food additives and ingredients used in food manufacturing across various countries is crucial for recognizing the diverse compounds involved. It enables the monitoring of reformulation processes and changes in the nutritional composition of products, particularly following the implementation of policies like front-of-package labeling.

These findings have implications for policymakers, public health professionals, and researchers. Understanding the impact of different identification scenarios on UPF prevalence and diagnostic test performance allows for more informed decision-making when establishing criteria for UPF classification. It also emphasizes the importance of considering the diverse nature of food categories, as certain products may require tailored criteria for accurate classification according to Nova food system.

In conclusion, this study enhances the understanding of UPF identification by highlighting the importance of detailed processes. We offer a replicable method that provides high sensitivity and specificity for identifying UPF, especially when traditional methods fall short due to unavailable ingredient list information. Our approach complements the classical method, facilitating safer decision-making in regulatory processes and the development of laws that clearly distinguish healthy and unhealthy foods based on the Nova classification. Our proposal can help the designing of policies that require precise definitions of UPFs and cannot afford moderate to high uncertainty. However, it should be noted that this is not the only method for UPF identification, and a comprehensive strategy should incorporate multiple approaches to ensure accuracy and effectiveness across different contexts.

## Data Availability

All data produced in the present work are contained in the manuscript

## Authors contribution

M.F.G and B.S.N. were responsible for the paper conceptualization, data curation, formal analysis, investigation, methodology and writing – original draft preparation. A.C.D. was responsible for the paper conceptualization, project administration, funding acquisition, and supervision, and contributed with data curation and interpretation, the paper methodology, and writing – review & editing., C.Z. contributed with the paper conceptualization, data curation and interpretation, paper methodology, and writing – review & editing. E.S. and L.G.B. contributed with formal analysis, paper methodology, and writing – review & editing. C.B. contributed with data curation, and paper conceptualization, methodology, supervision, and writing – review & editing.

