## Supplementary File 1; Supplementary File 2; Supplementary File 3;Supplementary File 4; Supplementary File 5; Supplementary File 6. for "Applying the Nova food classification to food product databases using discriminative ingredients: a methodological proposal"

**Supplementary material**

**Supplementary 1.** Food additives with potential cosmetic function. *Codex Alimentarius*, 2023.

| **Function** | **Food additive (INS)** |
| --- | --- |
| **Flavor enhancer** | Tartaric acid, L (+)- (334), Potassium chloride (508), Magnesium sulfate (518), Magnesium gluconate (580), Glutamic acid, L (+)- (620), Monosodium glutamate, L- (621), Monopotassium glutamate, L- (622), Calcium glutamate, di-L- (623), Monoammonium Glutamate, L- (624), Magnesium Glutamate, di-L- (625), Guanylic acid, 5'- (626), Disodium guanylate, 5'- (627), Dipotassium guanylate, 5'- (628), Calcium Guanylate, 5'- (629), Inosinic acid, 5'- (630), Disodium inosinate, 5'- (631), Potassium Inosinate, 5'- (632), Calcium Inosinate, 5'- (633), Calcium ribonucleotides, 5'- (634), Sodium ribonucleotides, 5'- (635), Maltol (636), Ethylmaltol (637), Acesulfame potassium (950), Aspartame (951), Sucralose (Trichlorogalactosaccharose) (955), Thaumatin (957), Neotamo (961), Erythritol (968), Advantage (969), Lipases (1104), Protease from Aspergillus orizae var. (1101 (i)), Papain (1101 (ii)), Bromelain (1101 (iii)) |
| **Color** | Tartrazine (102), Quinoline Amarillo (104), Ocaso Amarillo FCF (110), Carmines (120), Azorubin (Carmoisin) (122), Amaranth (123), Ponceau 4R (Rojo de cochinilla A) (124), Erythrosine (127), Red allura AC (129), Indigotine (Indigo Carmine) (132), Brilliant Blue FCF (133), Chlorophylls (140), Solid green FCF (143), Glossy Black (Black PN) (151), Brown HT (155), Purée red (162), Titanium dioxide (171), Tannic Acid (Tannins) (181), Curcumin (100 (i)), Riboflavin, synthetics (101 (i)), Riboflavin 5', sodium phosphate (101 (ii)), Riboflavin from Bacillus subtilis (101 (iii)), Chlorophylls, copper complexes (141 (i)), Chlorophyllines, copper complexes, potassium and sodium salts (141 (ii)), Caramel I – pure caramel (150a), Caramel II - sulphite caramel (150b), Caramel III - caramel with ammonia (150c), Caramel IV - caramel with ammonium sulfite (150d), Carotenes, beta-, synthetic (160a (i)), Carotenes, beta-, vegetables (160a (ii)), Carotenes, beta-, Blakeslea trispora- (160a (iii)), Dunaliella salina extract rich in beta-carotene (160 (aiv)), Annatto extracts, bixin base (160b (i)), Annatto extracts, norbixin base (160b (ii)), Chili extract (160c (ii)), Lycopene, synthetic (160d (i)), Lycopene, tomato (160d (ii)), Lycopene, Blakeslea trispora (160d (iii)), Carotenal, beta-apo-8'- (160e), Beta-apo-8'-carotenoic acid ethyl ester (160f), Tagetes erecta lutein (161b (i)), Lutein esters from Tagetes erecta (161b (iii)), Canthaxanthin (161g), Zeaxanthin, synthetic (161h (i)), Grape skin extract (163 (ii)), Calcium carbonate (170 (i)), Hierro oxide, black (172 (i)), Hierro oxide, red (172 (ii)), Hierro oxide, amarillo (172 (iii)) |
| **Emulsifier** | Tannic Acid (Tannins) (181), Sodium lactate (325), Potassium lactate (326), Alginic acid (400), Sodium alginate (401), Potassium alginate (402), Ammonium alginate (403), Propylene glycol alginate (405), Agar (406), Carrageenan (407), Algarrobo seed gum (410), Guar gum (412), Gum tragacanth (413), Gum Arabic (Acacia Gum) (414), Xanthan gum (415), Karaya gum (416), Ghatti gum (419), Octenylsuccinic acid (OSA) modified acacia gum (423), Harina konjac (425), Acacia gum (427), Polyoxyethylene stearate (8) (430), Polyoxyethylene stearate (40) (431), Polyoxyethylenated Sorbitan Monalaurate (20) (432), Polyoxyethylene sorbitan monooleate (20) (433), Polyoxyethylene sorbitan monopalmitate (20) (434), Polyoxyethylene sorbitan monostearate (20) (435), Polyoxyethylenated Sorbitan Tristearate (20) (436), Pectins (440), Ammonia sales of phosphatidic acid (442), Sucrose Isobutyrate Acetate (444), Methyl cellulose (461), Hydroxypropylcellulose (463), Hydroxypropyl methylcellulose (464), Methyl cellulose (465), Sodium Carboxymethyl Cellulose (Cellulose Gum) (466), Ethylhydroxyethylcellulose (467), Monoglycerides and Diglycerides of Fatty Acids (471), Fatty acid sucroesters (473), Sucroglycerides (474), Polyglyceride esters of fatty acids (475), Polyglyceride esters of interesterified ricinoleic acid (476), Propylene glycol esters of fatty acids (477), Oxidized soy oil with a thermal procedure interacted with monoglycerides and diglycerides of fatty acids (479), Sodium dioctyl sulfosuccinate (480), Stearyl citrate (484), Sorbitan Monostearate (491), Sorbitan Tristearate (492), Sorbitan monolaurate (493), Sorbitan Monooleate (494), Sorbitan Monopalmitate (495), Huesos phosphate (542), Beeswax (901), Candelilla wax (902), Lactitol (966), Xylitol (967), Polyvinylpyrrolidone (1201), Dextrins, toasted starch (1400), Acid-treated starch (1401), Almidón treated with alkali (1402), Blanched starch (1403), Oxidized starch (1404), Almidones treated with enzymes (1405), Monoalmidon phosphate (1410), Dialmidon Phosphate (1412), Phosphate Dialmidon Phosphate (1413), Acetylated Dialmidon Phosphate (1414), Starch Acetate (1420), Acetylated Dialmidon Adipate (1422), Hydroxypropyl starch (1440), Dialmidon Hydroxypropyl Phosphate (1442), Starch sodium octenyl succinate (1450), Oxidized acetylated starch (1451), Castor oil (1503), Triethyl citrate (1505), Triacetin (1518), Propylene glycol (1520), Polyethylene glycol (1521), Lecithin (322 (i)), Lecithin, partially hydrolyzed (322 (ii)), Sodium diacid citrate (331 (i)), Trisodium citrate (331 (iii)), Monosodium orthophosphate (339 (i)), Disodium hydrogen phosphate (339 (ii)), Trisodium phosphate (339 (iii)), Potassium diacid phosphate (340 (i)), Dipotassium hydrogen phosphate (340 (ii)), Tripotass phosphate (340 (iii)), Tricalcium phosphate (341 (iii)), Elaborate euchema seaweed (407a), Wood rosin glycerol esters (445 (iii)), Disodium diphosphate (450 (i)), Trisodium diphosphate (450 (ii)), Tetrasodium diphosphate (450 (iii)), Tetrapotassium diphosphate (450 (v)), Dicalcium diphosphate (450 (vi)), Calcium Diacid Diphosphate (450 (vii)), Pentasodium triphosphate (451 (i)), Pentapotassium triphosphate (451 (ii)), Sodium polyphosphate (452 (i)), Potassium polyphosphate (452 (ii)), Sodium and calcium polyphosphate (452 (iii)), Calcium polyphosphates (452 (iv)), Ammonium polyphosphates (452 (v)), Microcrystalline cellulose (Cellulose gel) (460 (i)), Cellulose in octopus (460 (ii)), Myristic, palmitic salt and stearic acids with ammonium, calcium, potassium and sodium (470 (i)), Oleic acid salt with calcium, potassium and sodium (470 (ii)), Magnesium stearate (470 (iii)), Acetic and fatty acid esters of glycerol (472a), Lactic and fatty acid esters of glycerol (472b), Citric and fatty acid esters of glycerol (472c), Diacetyl tartaric and fatty acid esters of glycerol (472e), Type I and II Sucrose Oligoesters (473a), Sodium stearoyl lactylate (481 (i)), Calcium stearoyl lactylate (482 (i)), Sodium and aluminum phosphate, acid (541 (i)), Aluminum and Sodium Phosphate, Basic (541 (ii)), Polydimethylsiloxane (900a), Maltitol (965 (i)), Maltitol jarab (965 (ii)), Quilaya extract, type 1 (999 (i)), Quilaya extract, type 2 (999 (ii)) |
| **Emulsifying salt** | Sodium lactate (325), Calcium lactate (327), Potassium and sodium tartrate, L (+)- (337), Tamarind seed polysaccharide (437), Sodium diacid citrate (331 (i)), Trisodium citrate (331 (iii)), Potassium Diacid Citrate (332 (i)), Tripotase citrate (332 (ii)), Tricalcium citrate (333 (iii)), Sodium tartrate, L (+)- (335 (ii)), Monosodium orthophosphate (339 (i)), Disodium hydrogen phosphate (339 (ii)), Tripotass phosphate (340 (iii)), Calcium diacid phosphate (341 (i)), Calcium hydrogen phosphate (341 (ii)), Tricalcium phosphate (341 (iii)), Magnesium Diacid Phosphate (343 (i)), Magnesium hydrogen phosphate (343 (ii)), Disodium diphosphate (450 (i)), Trisodium diphosphate (450 (ii)), Tetrasodium diphosphate (450 (iii)), Tetrapotassium diphosphate (450 (v)), Dicalcium diphosphate (450 (vi)), Calcium Diacid Diphosphate (450 (vii)), Pentasodium triphosphate (451 (i)), Pentapotassium triphosphate (451 (ii)), Sodium polyphosphate (452 (i)), Potassium polyphosphate (452 (ii)), Calcium polyphosphates (452 (iv)), Ammonium polyphosphates (452 (v)), Sodium carbonate (500 (i)), Sodium and aluminum phosphate, acid (541 (i)), Aluminum and Sodium Phosphate, Basic (541 (ii)) |
| **Sweetener** | Mannitol (421), Acesulfame potassium (950), Aspartame (951), Isomaltol (Hydrogenated Isomaltulose) (953), Sucralose (Trichlorogalactosaccharose) (955), Alitame (956), Thaumatin (957), Neotamo (961), Aspartame and Acesulfame Salt (962), Polyglycitol jarab (964), Lactitol (966), Xylitol (967), Erythritol (968), Advantage (969), Sorbitol (420 (i)), Sorbitol Jarab (420 (ii)), Cyclamic acid (952 (i)), Calcium cyclamate (952 (ii)), Sodium cyclamate (952 (iv)), Saccharin (954 (i)), Calcium saccharin (954 (ii)), Potassium saccharin (954 (iii)), Sodium saccharin (954 (iv)), Steviol glycosides from Stevia rebaudiana Bertoni (stevia glycosides from stevia) (960a), Rebaudioside A from multiple gene donors expressed in Yarrowialipolytica (960b (i)), Maltitol (965 (i)), Maltitol jarab (965 (ii)) |
| **Thickener** | Tannic Acid (Tannins) (181), Sodium lactate (325), Calcium lactate (327), Alginic acid (400), Sodium alginate (401), Potassium alginate (402), Ammonium alginate (403), Calcium alginate (404), Propylene glycol alginate (405), Agar (406), Carrageenan (407), Algarrobo seed gum (410), Guar gum (412), Gum tragacanth (413), Gum Arabic (Acacia Gum) (414), Xanthan gum (415), Karaya gum (416), Tara gum (417), Gellan gum (418), Ghatti gum (419), Mannitol (421), Glycerol (422), Curdlan (424), Harina konjac (425), Acacia gum (427), Tamarind seed polysaccharide (437), Pectins (440), Cyclodextrin, alpha- (457), Cyclodextrin, gamma- (458), Cyclodextrin, beta- (459), Methyl cellulose (461), Ethyl cellulose (462), Hydroxypropylcellulose (463), Hydroxypropyl methylcellulose (464), Methyl cellulose (465), Sodium Carboxymethyl Cellulose (Cellulose Gum) (466), Ethylhydroxyethylcellulose (467), Cross-linked Sodium Carboxymethyl Cellulose (Cross-linked Cellulose Gum) (468), Sodium carboxymethylcellulose, hydrolyzed by enzymes (Cellulose gum hydrolyzed by enzymes) (469), Potassium chloride (508), Calcium chloride (509), Sodium gluconate (576), Beeswax (901), Candelilla wax (902), Isomaltol (Hydrogenated Isomaltulose) (953), Lactitol (966), Xylitol (967), Polydextrose (1200), Polyvinylpyrrolidone (1201), Polyvinyl alcohol (1203), Pullulan (1204), Dextrins, toasted starch (1400), Acid-treated starch (1401), Almidón treated with alkali (1402), Blanched starch (1403), Oxidized starch (1404), Almidones treated with enzymes (1405), Monoalmidon phosphate (1410), Dialmidon Phosphate (1412), Phosphate Dialmidon Phosphate (1413), Acetylated Dialmidon Phosphate (1414), Starch Acetate (1420), Acetylated Dialmidon Adipate (1422), Hydroxypropyl starch (1440), Dialmidon Hydroxypropyl Phosphate (1442), Starch sodium octenyl succinate (1450), Oxidized acetylated starch (1451), Polyethylene glycol (1521), Monosodium orthophosphate (339 (i)), Disodium hydrogen phosphate (339 (ii)), Trisodium phosphate (339 (iii)), Potassium diacid phosphate (340 (i)), Dipotassium hydrogen phosphate (340 (ii)), Tripotass phosphate (340 (iii)), Calcium diacid phosphate (341 (i)), Calcium hydrogen phosphate (341 (ii)), Tricalcium phosphate (341 (iii)), Ammonium diacid phosphate (342 (i)), Diammonium hydrogen phosphate (342 (ii)), Magnesium Diacid Phosphate (343 (i)), Magnesium hydrogen phosphate (343 (ii)), Trimagnesium phosphate (343 (iii)), Elaborate euchema seaweed (407a), Sorbitol (420 (i)), Sorbitol Jarab (420 (ii)), Disodium diphosphate (450 (i)), Trisodium diphosphate (450 (ii)), Tetrasodium diphosphate (450 (iii)), Tetrapotassium diphosphate (450 (v)), Dicalcium diphosphate (450 (vi)), Pentasodium triphosphate (451 (i)), Pentapotassium triphosphate (451 (ii)), Sodium polyphosphate (452 (i)), Potassium polyphosphate (452 (ii)), Calcium polyphosphates (452 (iv)), Ammonium polyphosphates (452 (v)), Microcrystalline cellulose (Cellulose gel) (460 (i)), Cellulose in octopus (460 (ii)), Magnesium stearate (470 (iii)), Sodium carbonate (500 (i)), Sodium acid carbonate (500 (ii)), Sodium and aluminum phosphate, acid (541 (i)), Aluminum and Sodium Phosphate, Basic (541 (ii)), Baby powder (553 (iii)), Maltitol (965 (i)), Maltitol jarab (965 (ii)) |
| **Anti-foaming** | Carbon dioxide (290), Alginic acid (400), Sodium alginate (401), Potassium alginate (402), Ammonium alginate (403), Calcium alginate (404), Propylene glycol alginate (405), Xanthan gum (415), Hydroxypropylcellulose (463), Methyl cellulose (465), Fatty acid sucroesters (473), Nitrogen (941), Nitrous oxide (942), Microcrystalline cellulose (Cellulose gel) (460 (i)), Sodium stearoyl lactylate (481 (i)), Calcium stearoyl lactylate (482 (i)), Quilaya extract, type 1 (999 (i)), Quilaya extract, type 2 (999 (ii)) |
| **Bulking** | Sodium lactate (325), Alginic acid (400), Sodium alginate (401), Potassium alginate (402), Ammonium alginate (403), Calcium alginate (404), Propylene glycol alginate (405), Agar (406), Carrageenan (407), Gum Arabic (Acacia Gum) (414), Mannitol (421), Methyl cellulose (461), Ethyl cellulose (462), Hydroxypropyl methylcellulose (464), Sodium Carboxymethyl Cellulose (Cellulose Gum) (466), Carnauba wax (903), Isomaltol (Hydrogenated Isomaltulose) (953), Polydextrose (1200), Elaborate euchema seaweed (407 (a)), Sorbitol (420 (i)), Sorbitol Jarab (420 (ii)), Microcrystalline cellulose (Cellulose gel) (460 (i)), Cellulose in octopus (460 (ii)), Maltitol (965 (i)), Maltitol jarab (965 (ii)) |
| **Carbonating** | Carbon dioxide (290) |
| **Foaming** | Carbon dioxide (290), Carbon dioxide (290), Alginic acid (400), Sodium alginate (401), Potassium alginate (402), Ammonium alginate (403), Calcium alginate (404), Propylene glycol alginate (405), Xanthan gum (415), Hydroxypropylcellulose (463), Methyl cellulose (465), Fatty acid sucroesters (473), Nitrogen (941), Nitrous oxide (942), Microcrystalline cellulose (Cellulose gel) (460 (i)), Sodium stearoyl lactylate (481 (i)), Calcium stearoyl lactylate (482 (i)), Quilaya extract, type 1 (999 (i)), Quilaya extract, type 2 (999 (ii)) |
| **Gelling** | Alginic acid (400), Sodium alginate (401), Potassium alginate (402), Ammonium alginate (403), Calcium alginate (404), Propylene glycol alginate (405), Agar (406), Carrageenan (407), Tara gum (417), Gellan gum (418), Curdlan (424), Harina konjac (425), Acacia gum (427), Tamarind seed polysaccharide (437), Pectins (440), Sodium Carboxymethyl Cellulose (Cellulose Gum) (466), Elaborate euchema seaweed (407ª) |
| **Glazing agents** | Alginic acid (400), Sodium alginate (401), Potassium alginate (402), Ammonium alginate (403), Calcium alginate (404), Agar (406), Carrageenan (407), Gum Arabic (Acacia Gum) (414), Harina konjac (425), Pectins (440), Methyl cellulose (461), Ethyl cellulose (462), Hydroxypropylcellulose (463), Hydroxypropyl methylcellulose (464), Sodium Carboxymethyl Cellulose (Cellulose Gum) (466), Monoglycerides and Diglycerides of Fatty Acids (471), Fatty acid sucroesters (473), Beeswax (901), Candelilla wax (902), Carnauba wax (903), Shellac, bleached (904), Hydrogenated poly-1-decene (907), Isomaltol (Hydrogenated Isomaltulose) (953), Polydextrose (1200), Polyvinylpyrrolidone (1201), Polyvinyl alcohol (1203), Pullulan (1204), Basic methacrylate copolymer (CMB) (1205), Polyvinylalcohol (PVA)-Polyethylene Glycol graft copolymer (1209), Castor oil (1503), Propylene glycol (1520), Polyethylene glycol (1521), Elaborate euchema seaweed (407a), Microcrystalline cellulose (Cellulose gel) (460 (i)), Cellulose in octopus (460 (ii)), Type I and II Sucrose Oligoesters (473a), Baby powder (553 (iii)), Microcrystalline wax (905ci), High viscosity mineral oil (905d), Mineral oil, medium viscosity (905e) |

**Source:** Codex General Standard for Food Additives, 2023

**Supplementary 2.** Terms to identify flavorings.

| **Function** | **Terms** |
| --- | --- |
| **Flavor** | Flavoring, Natural flavoring, Natural flavor, Flavors identical to natural, Artificial flavoring, Essential oil |

**Source:** ANVISA, 2007

**Supplementary 3.** Food substances of rare or no culinary use.

| **Derivatives of added sugars** | **Derivatives of carbohydrates** | **Modified oils** | **Derivatives of**  **protein** | **Added Fiber** |
| --- | --- | --- | --- | --- |
| Inverted sugar  Dextrose  Frutose  Lactose  Glicose  Concentrated juice  Syrup | Dextrin  Maltodextrin  Modified starch | Interesterified vegetable oil  Interesterified fat  Hydrogenated oil  Hydrogenated vegetable oil  Hydrogenated vegetal fat  Fractioned oil  Fractioned fat | Concentrated protein  Protein isolate  Whey  Whey protein  Soy protein  Gluten  Casein  Gelatin  Pectin  Dairy product solids  Wheat gluten  Texturized soy protein  Mechanically separated meat | Soluble fiber  Insoluble fiber  Fiber |

**Source:** Monteiro, et al., 2019.

**Supplementary 4.** Description of the terms used in each scenario.

**Scenario 1 - food byproducts**

**Derivatives of added sugars:** inverted sugar; dextrose; frutose; lactose; glicose; concentrated juice; syrup

**Derivatives of carbohydrates:** dextrin; maltodextrin; modified starch

**Modified oils:** interesterified vegetable oil; interesterified fat; hydrogenated oil; hydrogenated vegetable oil; hydrogenated vegetal fat; fractioned oil; fractioned fat

**Derivatives of protein:** concentrated protein; protein isolate; whey; whey protein; soy protein; gluten; casein; gelatin; pectin; dairy product solids; wheat gluten; texturized soy protein; mechanically separated meat

**Added Fiber:** soluble fiber; insoluble fiber; fiber

**Scenario 2 - food additives (INS) or terms with sole cosmetic function**

**Flavor enhancer:** Papain (1101ii) ,Lipases (1104) ,Glutamic Acid, L(+)- (620) ,Monosodium Glutamate, L- (621) ,Monopotassium Glutamate, L- (622) ,Calcium Glutamate, Di-L- (623) ,Monoammonium Glutamate, L- (624) ,Magnesium Glutamate, Di-L- (625) ,Guanylic Acid, 5'- (626) ,Disodium Guanylate, 5'- (627) ,Dipotassium Guanylate, 5'- (628) ,Calcium Guanylate, 5'- (629) ,Inosinic Acid, 5'- (630) ,Disodium Inosinate, 5'- (631) ,Potassium Inosinate, 5'- (632) ,Calcium Inosinate, 5'- (633) ,Calcium Ribonucleotides, 5'- (634) ,Sodium Ribonucleotides, 5'- (635) ,Maltol (636) ,Ethylmaltol (637) ,Acesulfame Potassium (950) ,Aspartame (951) ,Sucralose (Trichlorogalactosaccharose) (955) ,Thaumatin (957)

**Glazing agents:** Shellac, Bleached (904) ,Mineral Oil, Medium Viscosity (905e), Hydrogenated Poly-1-Decene (907)

**Anti-foaming:** Microcrystalline Wax (905ci) , High Viscosity Mineral Oil (905d)

**Color:** Curcumin (100i) , Tartrazine (102) , Quinoline Amarillo (104) , Ocaso Amarillo FCF (110) , Carmines (120) , Azorubin (Carmoisin) (122) , Amaranth (123) , Ponceau 4R (Rojo De Cochinilla A) (124) , Erythrosine (127) , Red Allura AC (129) , Indigotine (Indigo Carmine) (132) , Brilliant Blue FCF (133) , Chlorophylls (140) , Chlorophylls, Copper Complexes (141i) , Chlorophyllines, Copper Complexes, Potassium And Sodium Salts (141ii) , Solid Green FCF (143) , Caramel I – Pure Caramel (150a) , Caramel II - Sulphite Caramel (150b) , Caramel III - Caramel With Ammonia (150c) , Caramel IV - Caramel With Ammonium Sulfite (150d) , Glossy Black (Black PN) (151) , Brown HT (155), Dunaliella Salina Extract Rich In Beta-Carotene (160aiv) , Annatto Extracts, Bixin Base (160bi) , Annatto Extracts, Norbixin Base (160bii) , Chili Extract (160cii) , Lycopene, Synthetic (160di) , Lycopene, Tomato (160dii) , Lycopene, Blakeslea Trispora (160diii) , Tagetes Erecta Lutein (161bi) , Lutein Esters From Tagetes Erecta (161biii) , Canthaxanthin (161g) , Zeaxanthin, Synthetic (161hi) , Purée Red (162) , Grape Skin Extract (163ii) , Titanium Dioxide (171) , Hierro Oxide, Black (172i) , Hierro Oxide, Red (172ii) , Hierro Oxide, Amarillo (172iii)

**Sweetener:** Cyclamic Acid (952i) , Calcium Cyclamate (952ii) , Sodium Cyclamate (952iv) , Saccharin (954i) , Calcium Saccharin (954ii) , Potassium Saccharin (954iii) , Sodium Saccharin (954iv) , Alitame (956) , Steviol Glycosides From Stevia Rebaudiana Bertoni (Stevia Glycosides From Stevia) (960a) , (960b) , Rebaudioside A From Multiple Gene Donors Expressed In Yarrowialipolytica (960bi) , (960c) , (960d) , Neotamo (961) , Aspartame And Acesulfame Salt (962) , Polyglycitol Jarab (964) , Lactitol (966) , Advantage (969)

**Emulsifier:** Octenylsuccinic Acid (OSA) Modified Acacia Gum (423) , Polyoxyethylene Stearate (8) (430) , Polyoxyethylene Stearate (40) (431) , Polyoxyethylene Sorbitan Monopalmitate (20) (434) , Ammonia Sales Of Phosphatidic Acid (442) , Sucroglycerides (474) , Polyglyceride Esters Of Interesterified Ricinoleic Acid (476) , Propylene Glycol Esters Of Fatty Acids (477) , Oxidized Soy Oil With A Thermal Procedure Interacted With Monoglycerides And Diglycerides Of Fatty Acids (479) , Sorbitan Monopalmitate (495) , Quilaya Extract, Type 1 (999i)

**Thickener:** Polyvinyl Alcohol (1203) Pullulan (1204)

**Foaming:** Quilaya Extract, Type 2 (999ii)

**Flavor:** Flavoring, Natural flavoring, Natural flavor, Flavors identical to natural, Artificial flavoring, Essential oil

**Scenario 3**

Combining the terms of scenarios 1 and 2

**Scenario 4 - food additive that can serve as an additive with cosmetic function**

**Flavor enhancer:** Tartaric acid, L (+)- (334), Potassium chloride (508), Magnesium sulfate (518), Magnesium gluconate (580), Glutamic acid, L (+)- (620), Monosodium glutamate, L- (621), Monopotassium glutamate, L- (622), Calcium glutamate, di-L- (623), Monoammonium Glutamate, L- (624), Magnesium Glutamate, di-L- (625), Guanylic acid, 5'- (626), Disodium guanylate, 5'- (627), Dipotassium guanylate, 5'- (628), Calcium Guanylate, 5'- (629), Inosinic acid, 5'- (630), Disodium inosinate, 5'- (631), Potassium Inosinate, 5'- (632), Calcium Inosinate, 5'- (633), Calcium ribonucleotides, 5'- (634), Sodium ribonucleotides, 5'- (635), Maltol (636), Ethylmaltol (637), Acesulfame potassium (950), Aspartame (951), Sucralose (Trichlorogalactosaccharose) (955), Thaumatin (957), Neotamo (961), Erythritol (968), Advantage (969), Lipases (1104), Protease from Aspergillus orizae var. (1101 (i)), Papain (1101 (ii)), Bromelain (1101 (iii))

**Color:** Tartrazine (102), Quinoline Amarillo (104), Ocaso Amarillo FCF (110), Carmines (120), Azorubin (Carmoisin) (122), Amaranth (123), Ponceau 4R (Rojo de cochinilla A) (124), Erythrosine (127), Red allura AC (129), Indigotine (Indigo Carmine) (132), Brilliant Blue FCF (133), Chlorophylls (140), Solid green FCF (143), Glossy Black (Black PN) (151), Brown HT (155), Purée red (162), Titanium dioxide (171), Tannic Acid (Tannins) (181), Curcumin (100 (i)), Riboflavin, synthetics (101 (i)), Riboflavin 5', sodium phosphate (101 (ii)), Riboflavin from Bacillus subtilis (101 (iii)), Chlorophylls, copper complexes (141 (i)), Chlorophyllines, copper complexes, potassium and sodium salts (141 (ii)), Caramel I – pure caramel (150a), Caramel II - sulphite caramel (150b), Caramel III - caramel with ammonia (150c), Caramel IV - caramel with ammonium sulfite (150d), Carotenes, beta-, synthetic (160a (i)), Carotenes, beta-, vegetables (160a (ii)), Carotenes, beta-, Blakeslea trispora- (160a (iii)), Dunaliella salina extract rich in beta-carotene (160 (aiv)), Annatto extracts, bixin base (160b (i)), Annatto extracts, norbixin base (160b (ii)), Chili extract (160c (ii)), Lycopene, synthetic (160d (i)), Lycopene, tomato (160d (ii)), Lycopene, Blakeslea trispora (160d (iii)), Carotenal, beta-apo-8'- (160e), Beta-apo-8'-carotenoic acid ethyl ester (160f), Tagetes erecta lutein (161b (i)), Lutein esters from Tagetes erecta (161b (iii)), Canthaxanthin (161g), Zeaxanthin, synthetic (161h (i)), Grape skin extract (163 (ii)), Calcium carbonate (170 (i)), Hierro oxide, black (172 (i)), Hierro oxide, red (172 (ii)), Hierro oxide, amarillo (172 (iii))

**Emulsifier:** Tannic Acid (Tannins) (181), Sodium lactate (325), Potassium lactate (326), Alginic acid (400), Sodium alginate (401), Potassium alginate (402), Ammonium alginate (403), Propylene glycol alginate (405), Agar (406), Carrageenan (407), Algarrobo seed gum (410), Guar gum (412), Gum tragacanth (413), Gum Arabic (Acacia Gum) (414), Xanthan gum (415), Karaya gum (416), Ghatti gum (419), Octenylsuccinic acid (OSA) modified acacia gum (423), Harina konjac (425), Acacia gum (427), Polyoxyethylene stearate (8) (430), Polyoxyethylene stearate (40) (431), Polyoxyethylenated Sorbitan Monalaurate (20) (432), Polyoxyethylene sorbitan monooleate (20) (433), Polyoxyethylene sorbitan monopalmitate (20) (434), Polyoxyethylene sorbitan monostearate (20) (435), Polyoxyethylenated Sorbitan Tristearate (20) (436), Pectins (440), Ammonia sales of phosphatidic acid (442), Sucrose Isobutyrate Acetate (444), Methyl cellulose (461), Hydroxypropylcellulose (463), Hydroxypropyl methylcellulose (464), Methyl cellulose (465), Sodium Carboxymethyl Cellulose (Cellulose Gum) (466), Ethylhydroxyethylcellulose (467), Monoglycerides and Diglycerides of Fatty Acids (471), Fatty acid sucroesters (473), Sucroglycerides (474), Polyglyceride esters of fatty acids (475), Polyglyceride esters of interesterified ricinoleic acid (476), Propylene glycol esters of fatty acids (477), Oxidized soy oil with a thermal procedure interacted with monoglycerides and diglycerides of fatty acids (479), Sodium dioctyl sulfosuccinate (480), Stearyl citrate (484), Sorbitan Monostearate (491), Sorbitan Tristearate (492), Sorbitan monolaurate (493), Sorbitan Monooleate (494), Sorbitan Monopalmitate (495), Huesos phosphate (542), Beeswax (901), Candelilla wax (902), Lactitol (966), Xylitol (967), Polyvinylpyrrolidone (1201), Dextrins, toasted starch (1400), Acid-treated starch (1401), Almidón treated with alkali (1402), Blanched starch (1403), Oxidized starch (1404), Almidones treated with enzymes (1405), Monoalmidon phosphate (1410), Dialmidon Phosphate (1412), Phosphate Dialmidon Phosphate (1413), Acetylated Dialmidon Phosphate (1414), Starch Acetate (1420), Acetylated Dialmidon Adipate (1422), Hydroxypropyl starch (1440), Dialmidon Hydroxypropyl Phosphate (1442), Starch sodium octenyl succinate (1450), Oxidized acetylated starch (1451), Castor oil (1503), Triethyl citrate (1505), Triacetin (1518), Propylene glycol (1520), Polyethylene glycol (1521), Lecithin (322 (i)), Lecithin, partially hydrolyzed (322 (ii)), Sodium diacid citrate (331 (i)), Trisodium citrate (331 (iii)), Monosodium orthophosphate (339 (i)), Disodium hydrogen phosphate (339 (ii)), Trisodium phosphate (339 (iii)), Potassium diacid phosphate (340 (i)), Dipotassium hydrogen phosphate (340 (ii)), Tripotass phosphate (340 (iii)), Tricalcium phosphate (341 (iii)), Elaborate euchema seaweed (407a), Wood rosin glycerol esters (445 (iii)), Disodium diphosphate (450 (i)), Trisodium diphosphate (450 (ii)), Tetrasodium diphosphate (450 (iii)), Tetrapotassium diphosphate (450 (v)), Dicalcium diphosphate (450 (vi)), Calcium Diacid Diphosphate (450 (vii)), Pentasodium triphosphate (451 (i)), Pentapotassium triphosphate (451 (ii)), Sodium polyphosphate (452 (i)), Potassium polyphosphate (452 (ii)), Sodium and calcium polyphosphate (452 (iii)), Calcium polyphosphates (452 (iv)), Ammonium polyphosphates (452 (v)), Microcrystalline cellulose (Cellulose gel) (460 (i)), Cellulose in octopus (460 (ii)), Myristic, palmitic salt and stearic acids with ammonium, calcium, potassium and sodium (470 (i)), Oleic acid salt with calcium, potassium and sodium (470 (ii)), Magnesium stearate (470 (iii)), Acetic and fatty acid esters of glycerol (472a), Lactic and fatty acid esters of glycerol (472b), Citric and fatty acid esters of glycerol (472c), Diacetyl tartaric and fatty acid esters of glycerol (472e), Type I and II Sucrose Oligoesters (473a), Sodium stearoyl lactylate (481 (i)), Calcium stearoyl lactylate (482 (i)), Sodium and aluminum phosphate, acid (541 (i)), Aluminum and Sodium Phosphate, Basic (541 (ii)), Polydimethylsiloxane (900a), Maltitol (965 (i)), Maltitol jarab (965 (ii)), Quilaya extract, type 1 (999 (i)), Quilaya extract, type 2 (999 (ii))

**Emulsifying salt:** Sodium lactate (325), Calcium lactate (327), Potassium and sodium tartrate, L (+)- (337), Tamarind seed polysaccharide (437), Sodium diacid citrate (331 (i)), Trisodium citrate (331 (iii)), Potassium Diacid Citrate (332 (i)), Tripotase citrate (332 (ii)), Tricalcium citrate (333 (iii)), Sodium tartrate, L (+)- (335 (ii)), Monosodium orthophosphate (339 (i)), Disodium hydrogen phosphate (339 (ii)), Tripotass phosphate (340 (iii)), Calcium diacid phosphate (341 (i)), Calcium hydrogen phosphate (341 (ii)), Tricalcium phosphate (341 (iii)), Magnesium Diacid Phosphate (343 (i)), Magnesium hydrogen phosphate (343 (ii)), Disodium diphosphate (450 (i)), Trisodium diphosphate (450 (ii)), Tetrasodium diphosphate (450 (iii)), Tetrapotassium diphosphate (450 (v)), Dicalcium diphosphate (450 (vi)), Calcium Diacid Diphosphate (450 (vii)), Pentasodium triphosphate (451 (i)), Pentapotassium triphosphate (451 (ii)), Sodium polyphosphate (452 (i)), Potassium polyphosphate (452 (ii)), Calcium polyphosphates (452 (iv)), Ammonium polyphosphates (452 (v)), Sodium carbonate (500 (i)), Sodium and aluminum phosphate, acid (541 (i)), Aluminum and Sodium Phosphate, Basic (541 (ii))

**Sweetener:** Mannitol (421), Acesulfame potassium (950), Aspartame (951), Isomaltol (Hydrogenated Isomaltulose) (953), Sucralose (Trichlorogalactosaccharose) (955), Alitame (956), Thaumatin (957), Neotamo (961), Aspartame and Acesulfame Salt (962), Polyglycitol jarab (964), Lactitol (966), Xylitol (967), Erythritol (968), Advantage (969), Sorbitol (420 (i)), Sorbitol Jarab (420 (ii)), Cyclamic acid (952 (i)), Calcium cyclamate (952 (ii)), Sodium cyclamate (952 (iv)), Saccharin (954 (i)), Calcium saccharin (954 (ii)), Potassium saccharin (954 (iii)), Sodium saccharin (954 (iv)), Steviol glycosides from Stevia rebaudiana Bertoni (stevia glycosides from stevia) (960a), Rebaudioside A from multiple gene donors expressed in Yarrowialipolytica (960b (i)), Maltitol (965 (i)), Maltitol jarab (965 (ii))

**Thickener:** Tannic Acid (Tannins) (181), Sodium lactate (325), Calcium lactate (327), Alginic acid (400), Sodium alginate (401), Potassium alginate (402), Ammonium alginate (403), Calcium alginate (404), Propylene glycol alginate (405), Agar (406), Carrageenan (407), Algarrobo seed gum (410), Guar gum (412), Gum tragacanth (413), Gum Arabic (Acacia Gum) (414), Xanthan gum (415), Karaya gum (416), Tara gum (417), Gellan gum (418), Ghatti gum (419), Mannitol (421), Glycerol (422), Curdlan (424), Harina konjac (425), Acacia gum (427), Tamarind seed polysaccharide (437), Pectins (440), Cyclodextrin, alpha- (457), Cyclodextrin, gamma- (458), Cyclodextrin, beta- (459), Methyl cellulose (461), Ethyl cellulose (462), Hydroxypropylcellulose (463), Hydroxypropyl methylcellulose (464), Methyl cellulose (465), Sodium Carboxymethyl Cellulose (Cellulose Gum) (466), Ethylhydroxyethylcellulose (467), Cross-linked Sodium Carboxymethyl Cellulose (Cross-linked Cellulose Gum) (468), Sodium carboxymethylcellulose, hydrolyzed by enzymes (Cellulose gum hydrolyzed by enzymes) (469), Potassium chloride (508), Calcium chloride (509), Sodium gluconate (576), Beeswax (901), Candelilla wax (902), Isomaltol (Hydrogenated Isomaltulose) (953), Lactitol (966), Xylitol (967), Polydextrose (1200), Polyvinylpyrrolidone (1201), Polyvinyl alcohol (1203), Pullulan (1204), Dextrins, toasted starch (1400), Acid-treated starch (1401), Almidón treated with alkali (1402), Blanched starch (1403), Oxidized starch (1404), Almidones treated with enzymes (1405), Monoalmidon phosphate (1410), Dialmidon Phosphate (1412), Phosphate Dialmidon Phosphate (1413), Acetylated Dialmidon Phosphate (1414), Starch Acetate (1420), Acetylated Dialmidon Adipate (1422), Hydroxypropyl starch (1440), Dialmidon Hydroxypropyl Phosphate (1442), Starch sodium octenyl succinate (1450), Oxidized acetylated starch (1451), Polyethylene glycol (1521), Monosodium orthophosphate (339 (i)), Disodium hydrogen phosphate (339 (ii)), Trisodium phosphate (339 (iii)), Potassium diacid phosphate (340 (i)), Dipotassium hydrogen phosphate (340 (ii)), Tripotass phosphate (340 (iii)), Calcium diacid phosphate (341 (i)), Calcium hydrogen phosphate (341 (ii)), Tricalcium phosphate (341 (iii)), Ammonium diacid phosphate (342 (i)), Diammonium hydrogen phosphate (342 (ii)), Magnesium Diacid Phosphate (343 (i)), Magnesium hydrogen phosphate (343 (ii)), Trimagnesium phosphate (343 (iii)), Elaborate euchema seaweed (407a), Sorbitol (420 (i)), Sorbitol Jarab (420 (ii)), Disodium diphosphate (450 (i)), Trisodium diphosphate (450 (ii)), Tetrasodium diphosphate (450 (iii)), Tetrapotassium diphosphate (450 (v)), Dicalcium diphosphate (450 (vi)), Pentasodium triphosphate (451 (i)), Pentapotassium triphosphate (451 (ii)), Sodium polyphosphate (452 (i)), Potassium polyphosphate (452 (ii)), Calcium polyphosphates (452 (iv)), Ammonium polyphosphates (452 (v)), Microcrystalline cellulose (Cellulose gel) (460 (i)), Cellulose in octopus (460 (ii)), Magnesium stearate (470 (iii)), Sodium carbonate (500 (i)), Sodium acid carbonate (500 (ii)), Sodium and aluminum phosphate, acid (541 (i)), Aluminum and Sodium Phosphate, Basic (541 (ii)), Baby powder (553 (iii)), Maltitol (965 (i)), Maltitol jarab (965 (ii))

**Anti-foaming:** Carbon dioxide (290), Alginic acid (400), Sodium alginate (401), Potassium alginate (402), Ammonium alginate (403), Calcium alginate (404), Propylene glycol alginate (405), Xanthan gum (415), Hydroxypropylcellulose (463), Methyl cellulose (465), Fatty acid sucroesters (473), Nitrogen (941), Nitrous oxide (942), Microcrystalline cellulose (Cellulose gel) (460 (i)), Sodium stearoyl lactylate (481 (i)), Calcium stearoyl lactylate (482 (i)), Quilaya extract, type 1 (999 (i)), Quilaya extract, type 2 (999 (ii))

**Bulking:** Sodium lactate (325), Alginic acid (400), Sodium alginate (401), Potassium alginate (402), Ammonium alginate (403), Calcium alginate (404), Propylene glycol alginate (405), Agar (406), Carrageenan (407), Gum Arabic (Acacia Gum) (414), Mannitol (421), Methyl cellulose (461), Ethyl cellulose (462), Hydroxypropyl methylcellulose (464), Sodium Carboxymethyl Cellulose (Cellulose Gum) (466), Carnauba wax (903), Isomaltol (Hydrogenated Isomaltulose) (953), Polydextrose (1200), Elaborate euchema seaweed (407 (a)), Sorbitol (420 (i)), Sorbitol Jarab (420 (ii)), Microcrystalline cellulose (Cellulose gel) (460 (i)), Cellulose in octopus (460 (ii)), Maltitol (965 (i)), Maltitol jarab (965 (ii))

**Carbonating:** Carbon dioxide (290)

**Foaming:** Carbon dioxide (290), Carbon dioxide (290), Alginic acid (400), Sodium alginate (401), Potassium alginate (402), Ammonium alginate (403), Calcium alginate (404), Propylene glycol alginate (405), Xanthan gum (415), Hydroxypropylcellulose (463), Methyl cellulose (465), Fatty acid sucroesters (473), Nitrogen (941), Nitrous oxide (942), Microcrystalline cellulose (Cellulose gel) (460 (i)), Sodium stearoyl lactylate (481 (i)), Calcium stearoyl lactylate (482 (i)), Quilaya extract, type 1 (999 (i)), Quilaya extract, type 2 (999 (ii))

**Gelling:** Alginic acid (400), Sodium alginate (401), Potassium alginate (402), Ammonium alginate (403), Calcium alginate (404), Propylene glycol alginate (405), Agar (406), Carrageenan (407), Tara gum (417), Gellan gum (418), Curdlan (424), Harina konjac (425), Acacia gum (427), Tamarind seed polysaccharide (437), Pectins (440), Sodium Carboxymethyl Cellulose (Cellulose Gum) (466), Elaborate euchema seaweed (407ª)

**Glazing agents:** Alginic acid (400), Sodium alginate (401), Potassium alginate (402), Ammonium alginate (403), Calcium alginate (404), Agar (406), Carrageenan (407), Gum Arabic (Acacia Gum) (414), Harina konjac (425), Pectins (440), Methyl cellulose (461), Ethyl cellulose (462), Hydroxypropylcellulose (463), Hydroxypropyl methylcellulose (464), Sodium Carboxymethyl Cellulose (Cellulose Gum) (466), Monoglycerides and Diglycerides of Fatty Acids (471), Fatty acid sucroesters (473), Beeswax (901), Candelilla wax (902), Carnauba wax (903), Shellac, bleached (904), Hydrogenated poly-1-decene (907), Isomaltol (Hydrogenated Isomaltulose) (953), Polydextrose (1200), Polyvinylpyrrolidone (1201), Polyvinyl alcohol (1203), Pullulan (1204), Basic methacrylate copolymer (CMB) (1205), Polyvinylalcohol (PVA)-Polyethylene Glycol graft copolymer (1209), Castor oil (1503), Propylene glycol (1520), Polyethylene glycol (1521), Elaborate euchema seaweed (407a), Microcrystalline cellulose (Cellulose gel) (460 (i)), Cellulose in octopus (460 (ii)), Type I and II Sucrose Oligoesters (473a), Baby powder (553 (iii)), Microcrystalline wax (905ci), High viscosity mineral oil (905d), Mineral oil, medium viscosity (905e)

**Flavor:** Flavoring, Natural flavoring, Natural flavor, Flavors identical to natural, Artificial flavoring, Essential oil

**Scenario 5**

Combining the terms of scenarios 1 and 4

**Supplementary 5.** Vitamins and minerals that can serve as an additive of cosmetic function, 2021 *Codex Alimentarius.*

| **Function** | **Food additive (INS)** |
| --- | --- |
| **Color** | Riboflavin, synthetics (101i) ,Riboflavin 5', sodium phosphate (101ii) ,Riboflavin from Bacillus subtilis (101iii) , Carotenes, beta-, synthetic (160ai) ,Carotenes, beta-, vegetables (160aii) ,Carotenes, beta-, Blakeslea trispora- (160aiii), Carotenal, beta-apo-8'- (160e) ,Beta-apo-8'-carotenoic acid ethyl ester (160f); Calcium carbonate (170 (i)) |

**Source:** Codex General Standard for Food Additives, 2023.

**Supplementary 6.** Proportions of UPF and test diagnostics by scenarios with inclusion of vitamins and minerals

| **Scenarios** | **% (CI 95%)*** | **p-value**** | **Sensitivity** | **Specificity** | **Positive predictive value** | **Negative predictive value** |
| --- | --- | --- | --- | --- | --- | --- |
| **Scenario 2** | 55,0 (54,00; 55,96) | p≥0.005 | 71.3% | 83.9% | 91.3% | 55.1% |
| **Scenario 2 + vitamins and minerals** | 55,1 (54,16; 56,12) | - | 71.5% | 83.9% | 91.4% | 55.3% |
| **Scenario 3** | 65,0 (64,08; 65,96) | p≥0.005 | 82.6% | 76.8% | 89.4% | 64.9% |
| **Scenario 3 + vitamins and minerals** | 65,1 (64,11; 65,99) | - | 82.6% | 76.8% | 89.4% | 64.9% |

*Confidence Interval 95%; **Proportion test; Scenario 2: Presence of at least one food additive with sole cosmetic function; Scenario 3: Presence of at least one food byproduct or a food additive with sole cosmetic function.
